# Cognition in Whitehall II ’SuperAgers’: An Analysis of Dementia Risk Factors

**DOI:** 10.1101/2025.08.12.25333490

**Authors:** Philippa Watson, Omid Ebrahimi, John Gallacher, Sarah Bauermeister

## Abstract

While many experience cognitive decline, a subset called SuperAgers can reach late life with cognitive functioning comparable to those decades younger. We aimed to categorise whether dementia risk factors may be responsible for SuperAger status in old age. SuperAgers were identified in the UK Whitehall II cohort (n = 3513, 25.88% female) as participants 65 years or older who scored above the sex specific mean of mid-life individuals on a test of verbal memory and who scored within one standard deviation above the mean on a test of fluency in older age. Logistic regression was used to examine associations between late-life dementia risk factors and SuperAger status. Higher education (OR = 1.23, SE = 0.05, 95% CI = [1.10, 1.36], *p* < 0.001), female sex (OR = 1.30, SE = 0.09, 95% CI = [1.10, 1.55], *p* = 0.003), and lower age (OR = 0.67, SE = 0.04, 95% CI = [0.61, 0.73], *p* < 0.001) were associated with increased odds of SuperAger status. Absence of hypertension (OR = 0.76, SE = 0.08, 95% CI = [0.64, 0.89], *p* < 0.001), lower depressive symptoms (OR = 0.99, SE = 0.01, 95% CI = [0.98, 1.00], *p* = 0.05), higher fruit and vegetable intake (OR = 1.08, SE = 0.03, 95% CI = [1.02, 1.15], *p* = 0.01), and greater social activity engagement (OR = 1.03, SE = 0.01, 95% CI = [1.01, 1.06], *p* = 0.02) were also associated with higher odds. These findings suggest hypertension, depressive symptoms, diet, and social engagement show promising but modest associations with SuperAger status, however, they do not fully explain exceptional cognitive ageing.

**Author summary:** While many people experience cognitive decline as they age, a subset of older adults, known as “SuperAgers”, reach later life with cognitive functioning comparable to people decades younger. SuperAgers appear to resist age-related cognitive decline and can, for example, perform as well as or better than healthy adults 20–30 years younger on tests of memory. We aimed to investigate whether established dementia risk factors are associated with SuperAger status in older age. SuperAgers were identified in the UK Whitehall II cohort as participants aged 65 years or older who performed above the sex-specific mean of mid-life individuals on a test of verbal memory and within one standard deviation of the mean on a test of verbal fluency in older age. We examined associations between late-life dementia risk factors and SuperAger status. We found that the absence of hypertension, lower depression scores, greater fruit and vegetable intake, and higher social engagement were associated with an increased likelihood of being a SuperAger, although these associations were modest. Our study provides new insights into factors associated with exceptional cognitive ageing and suggests that established late-life dementia risk factors may contribute to, but do not fully explain, why some individuals maintain exceptionally high cognitive function into older age.

## Introduction

The global population of older adults (65+ years) is steadily increasing. In the UK, this demographic currently represents approximately 19% of the population, with this figure predicted to rise to 27% by 2072 (Office for National Statistics, 2024). Older individuals age-related cognitive change or decline is highly heterogeneous. While extensive research has examined cognitive impairment in this population, significantly less attention has been given to the other end of the spectrum: older adults exhibiting exceptional cognitive abilities (Petersen et al., 2014; Yu et al., 2020).

Although cognitive impairment is readily conceptualized through diagnoses like Alzheimer’s disease (AD) or mild cognitive impairment (MCI), defining ‘successful cognitive ageing’ has proven more challenging (Martin et al., 2015; Rowe & Kahn, 2015). One prominent conceptualization focuses on ‘SuperAgers’, defined as a person who appears to resist age-related cognitive decline and instead exhibits, for example, superior memory performance by scoring as high or higher as healthy adults 20–30 years younger (Garo-Pascual et al., 2023; Harrison et al., 2012; Maccora et al., 2021). Studies indicate that SuperAgers exhibit cortical preservation in specific brain regions associated with resilience (de Godoy et al., 2021). Notably, SuperAgers demonstrate greater cortical thickness in the hippocampus (Dekhtyar et al., 2017; Harrison et al., 2018; Sun et al., 2016; Yang et al., 2016), anterior cingulate cortex (Gefen et al., 2015; Harrison et al., 2012; Sun et al., 2016), medial prefrontal cortex (Harrison et al., 2018; Sun et al., 2016; Yang et al., 2016) (structures within the default mode network linked to episodic memory), and the insula (de Godoy et al., 2021; Harrison et al., 2018; Sun et al., 2016; Yang et al., 2016) (a salience network component involved in attention and executive processes during encoding and retrieval). Furthermore, research shows SuperAgers are less likely to carry an APOE-E4 allele and have reduced AD pathology frequency (Rogalski et al., 2013). Collectively, these findings suggest a distinctive neurobiological profile in SuperAgers, contrasting with the typical pattern of age-related cortical atrophy that begins in young adulthood and progresses throughout life (Cook Maher et al., 2022; Fotenos et al., 2005; Resnick et al., 2003).

While the neuroanatomical characteristics of SuperAgers are relatively well-documented, the potential factors (e.g., lifestyle, environmental) contributing to this status remain less explored. Identifying such factors is crucial, as they could provide insights into resilience mechanisms, inform strategies to prevent cognitive decline, and reduce the public health burden of cognitive decline and related disorders (e.g. dementia) (Anstey & Christensen, 2000; Burke et al., 2019).

Cumulative epidemiological evidence strongly links a healthy lifestyle to preserved cognitive health in later life. It is estimated that as much as 40% of worldwide dementia cases could be prevented if a healthy lifestyle is adhered to (Livingston et al., 2024). Specific risk factors such as physical activity, smoking tobacco, consuming alcohol, sleep habits, and social engagement have been intensively studied with accumulating evidence linking them to cognitive impairment and dementia risk (Baumgart et al., 2015; Lourida et al., 2019; Peters et al., 2019; Sabia et al., 2012). For example, a recent systematic review examining the role of modifiable risk factors on cognitive health and dementia in old age found that healthy diet, leisure and physical activity participation protected against cognitive decline (Ye et al., 2023). Given these links between risk factors and cognitive outcomes across the spectrum from impairment to healthy ageing, it is compelling to investigate whether similar factors contribute to the exceptional cognitive preservation observed in SuperAgers.

Preliminary research has begun exploring risk factors engagement in SuperAgers. Hermansen and colleagues (2024), for instance, found in a study investigating Cognitively High-Performing Oldest Old individuals (CHPOOI) in two Danish cohorts, that CHPOOI individuals had higher physical activity engagement alongside lower depression symptomology. However, research in this area remains limited with further studies needed to clarify these relationships.

Building on existing research, this paper aims to identify whether dementia risk factors are associated with superior cognitive ageing (i.e., SuperAger status) in a sample of healthy older adults from the Whitehall II cohort. This was assessed by simultaneously examining multiple dementia risk factors and their relationship to SuperAger status. We hypothesised that factors associated with prevention against dementia and healthy ageing (e.g., physical activity, mental health, social engagement) may be significantly stronger in the SuperAger group than in the healthy cognitive control group.

## Material and Methods

### Study design

Participants were drawn from the Whitehall II study, a longitudinal cohort study targeting British civil servants (3,413 women and 6,895 men) from various employment grades across 20 civil service departments. With a participation rate of 73%, the baseline cohort (1985–88) consisted of 10,308 civil servants aged 35–55 years. Assessments have alternated between a postal questionnaire only and a postal questionnaire and clinical examination. Written informed consent was obtained from cohort members before participation and ethical approval was granted by the University College London Ethics Committee. Further details about the cohort can be found elsewhere (Marmot & Brunner, 2005). To date there have been 12 waves of data collection. The present study uses data from Phase 1 (1985-88), Phase 5 (1997-1999) and Phase 12 (2015-2016) of Whitehall II study data collection all of which included a clinical examination. Seventy-eight percent of eligible Phase 1 respondents participated in Phase 5 (n = 7,870), and 67% participated in Phase 12 (n = 5,632). The present study was a secondary data analysis, with all participants from Whitehall II providing written informed consent (Marmot & Brunner, 2005), Whitehall II received ethics approval from the London-Harrow Research Ethics Committee, REC Ref 85/0938 and the Scotland A REC, REC Ref 16/SS/0003.

The median length of follow-up from Phase 5 to Phase 12 was 18 years; 2877 individuals died between study recruitment and phase 12. The analyses for the present study were based on 3513 participants who were 65 years or older at Phase 12, had no missing data on any of the study’s cognitive variables and met the classification for SuperAger or cognitively normal, see Fig. 1 below.

**Fig 1.**
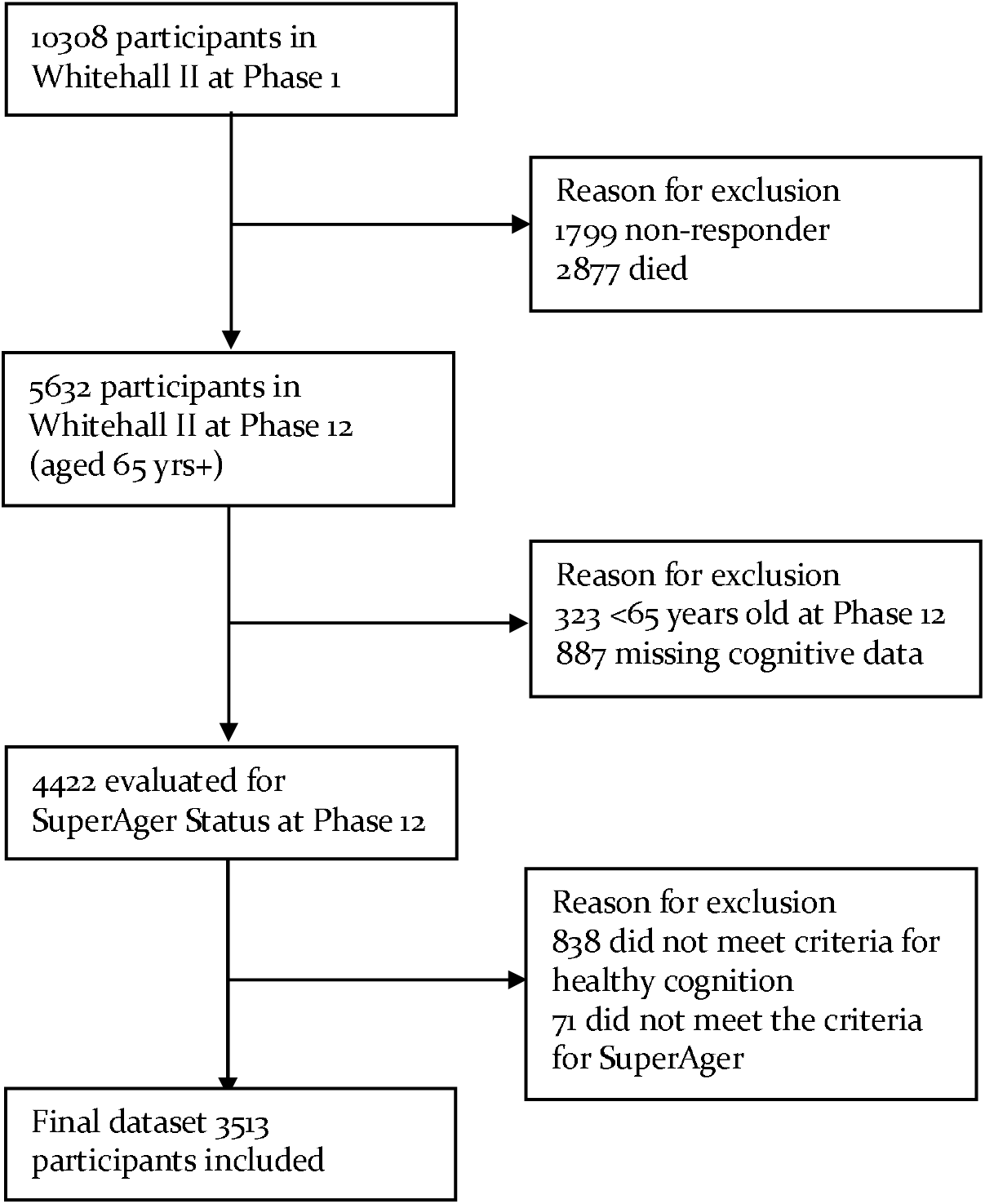
Attrition figure of study participants and selection criteria. No information was available on attrition from phase 5 to 12, therefore phase 1 to 12 attrition is reported.

### Study Variables

#### SuperAger Status

Cognitive data was examined at Phases 5 and 12. Phase 5 was used because it was the earliest point with relevant cognitive data, while Phase 12 was chosen as the most recent available point to maximise the time gap between assessments (18 years). Verbal memory was assessed with a 20-word free recall task, whereby participants were presented with a list of 20 one or two syllable words and then were asked to recall as many as possible within 2 minutes. Fluency was assessed with a 60-second written naming task of words beginning with the letter “S” (Borkowski et al., 1967). All 4,422 subjects were classified into three groups: SuperAgers, cognitively normal agers (CNs), and adults with cognitive impairment (CIs). The CI group were defined as participants who scored in the bottom tercile on a test of verbal memory in phase 12 (n = 838) and were excluded from the analysis given the present study focus on investigating determinates for cognitive health. In this study, SuperAgers were defined by age and stringent neuropsychological performance criteria, following criteria from the Northwestern SuperAging program (Harrison et al., 2012). Specifically, SuperAgers were those who had a verbal memory score at phase 12 that was above the sex specific phase 5 mean score, and had phase 12 fluency scores within one standard deviation of the sex specific fluency score mean (coded ‘1’). CNs had a verbal memory score at phase 12 within one standard deviation of the sex specific mean for phase 12 and had phase 12 fluency scores within 1.5 standard deviations of the sex specific fluency score mean (coded ‘0’).

#### Predictor Variables

We assessed 17 risk factors for dementia based on Lancet and existing literature (e.g., Dintica & Yaffe, 2022; Livingston et al., 2024; Mukadam et al., 2020; Mukadam et al., 2024), examining their association with SuperAger status. All variables were examined at phase 12 with the exception of education and fathers social class, which were evaluated at baseline. Variables were grouped into demographic, biomedical, lifestyle, and psychosocial.

Demographic variables included education, father’s social class, and sex. Education was measured by examining years of education split into three categories, up to 16 years, 17-18 years, and 19+ years. Father’s (occupational) social class was assessed at age 10-11 and classified using the Registrar-General’s Social Classes into the following groups: I (professional), II (managerial and technical), IIIN (skilled non-manual), IIIM (skilled manual), IV (partly skilled), and V (unskilled).

Biomedical variables derived from medical records included hypertension (systolic/diastolic blood pressure ≥140/90 mm Hg or use of antihypertensive drugs), cardiovascular disease (examining coronary heart disease and stroke), and diabetes (coded as present ‘1’/absent ‘0’). Self-reported number of medications calculated by summing number of current medications used, BMI calculated as weight in kilograms divided by the square of height in centimetres were also included, sight coded as poor sight (coded poor ‘1’/excellent-fair ‘0’), and hearing coded as poor hearing (coded poor ‘1’, excellent-fair ‘0’).

Lifestyle variables examined included sleep quality, alcohol, physical activity, smoking history, diet, and leisure activity engagement. Sleep quality was assessed using the four-item Jenkins sleep questionnaire (Jenkins et al., 1988) a questionnaire assessing how often participants had experienced sleep disturbances in the past month, with all items on a 6-item scale from 1 being not at all to 6 being 22-31 days. Alcohol was measured by examining the standard alcoholic drinks consumed per week, and smoking history was split into never smokers and current or former smokers. Physical activity was assessed via a self-report questionnaire that reflected the Minnesota leisure-time physical activity questionnaire (Richardson et al., 1994), with participants reporting the frequency and duration of mild, moderate, and vigorous activities. These data were used to derive metabolic equivalent of task (MET) values, with moderate-to-vigorous physical activity classified according to established guidelines (Ainsworth et al., 2000; Sabia et al., 2017). Diet was measured by self-reported vegetable and fruit intake on a scale of 1 (seldom consume) to 9 (4+ daily). Leisure activity engagement was assessed across 13 activities and summarised into social and individual activity scores (Singh-Manoux et al., 2003). An activity was classified as social if they required engagement with others, and individual if it did not, for further details see Sing-Manous and colleagues (2003).

Psychosocial variables included the Centre for Epidemiologic Studies Depression Scale score (CES-D; Radloff, 1977). The CES-D encompasses 20 items, rated on a 4-point scale (from 0: rarely or none of the time to 3 most or all of the time), with scores ranging from 0 to 60, where higher scores indicate a more pronounced level of depression.

### Statistical Analyses

All analyses were carried out in the Dementias Platform UK (DPUK) Data Portal (Bauermeister et al., 2020) and analysed using STATA SE 17.0 47 and R version 4.5.2. Participants’ characteristics at the most recent wave (phase 12) were described using means (Standard Errors) and counts (%) split by SuperAger group. Statistical comparisons were also performed between groups, with chi-squared tests conducted for categorical variables and *t*-tests for continuous variables to determine associations between potential risk factors and SuperAger status.

The proportion of missing data across variables ranged from 0% to 32.9%. The highest missingness was observed in vision loss (32.9%), followed by alcohol consumption (27.4%), smoking status (10.5%), individualised activity engagement (7.2%), and Jenkin’s sleep problem score (6.2%). Most other variables had less than 6% missing data. At the participant level, of the 3513 individuals 1770 had missingness in at least one variable. Of those with missing data, 79.2% were missing data in only one or two variables, with a further 15.5% missing three to four variables, this indicating generally low levels of missingness.

Missing data was addressed using multiple imputation by chained equations (MICE) using the mice package in R (White et al., 2011). Thirty imputed datasets were generated with 20 iterations per dataset. Continuous variables were imputed using predictive mean matching, binary variables using logistic regression, and categorical variables using proportional odds logistic regression. The imputed model included all variables used in the analysis with participant identifiers excluded from imputation and predictor matrices. Estimates from regression analyses were pooled across imputed datasets using Rubin’s rules (Rubin, 2018).

To investigate associations between potential risk factors and SuperAger status a binary logistic regression was conducted. Any variable which was observed to have a significant association (*p* < 0.05) in the descriptive statistics were input into the logistic regression. Pooled estimates across imputed datasets were obtained, and results were reported as odd ratios (ORs) with 95% confidence intervals (CIs) and standard errors (SEs).

## Results

The sample included 1137 SuperAgers and 2376 CNs, who met the inclusion criteria and were all included in the present analysis. Descriptive statistics of the participants are presented in Table 2. SuperAgers were more likely to be female (48.07% female SuperAgers vs 40.01% male SuperAgers, *p* < 0.001) and had higher educational levels than CNs (*p* < 0.001). Several potential risk factors were observed to be associated with being a SuperAger. Higher social activity participation, higher individual activity participation, lower presence of hypertension, less total medications used, lower BMI, and lower depression score were significant predictors of being a SuperAger. All variables that showed significant univariate associations with SuperAger status were brought forward to the multiple logistic regression model.

**Table 2.** Sample descriptives at selected baseline (Phase 12) for SuperAgers and CNs.

| Variable |  | CN<br>M(SD) or % | SuperAger<br>M(SD) or % | p value |
| --- | --- | --- | --- | --- |
| Demographic |  |  |  |  |
| Sex (% female) |  | 26.6% | 29.6% | 0.08 |
| Age group | 65-69 yrs | 38.0% | 56.6% | <0.001*** |
|  | 70-74 yrs | 29.7% | 27.6% |  |
|  | 75-79 yrs | 20.0% | 11.3% |  |
|  | 80-85 yrs | 12.0% | 4.0% |  |
|  | 85+ yrs | 0.3% | 0.5% |  |
| Education Level | Up to 16 | 29.8% | 18.9% | <0.001*** |
|  | 17-18 | 25.3% | 25.9% |  |
|  | Over 18 | 44.9% | 55.2% |  |
| Father's social class | Class I | 11.1% | 11.5% | 0.003** |
|  | Class II | 31.4% | 37.1% |  |
|  | Class III Non-Manual | 16.6% | 15.5% |  |
|  | Class III Manual | 31.8% | 27.2% |  |
|  | Class IV | 5.8% | 7.2% |  |
|  | Class V | 3.3% | 1.4% |  |
| Biomedical |  |  |  |  |
| Total BMI |  | 26.95 (4.44) | 26.47 (4.37) | 0.002** |
| Total medications |  | 3.52 (2.82) | 3.04 (2.55) | <0.001*** |
| Hypertension (%) |  | 59.0% | 47.8% | <0.001*** |
| Diabetes (%) |  | 9.5% | 7.3% | 0.04* |
| Cardiovascular diseases (%) |  | 4.8% | 3.5% | 0.09 |
| Hearing loss (%) |  | 1.1% | 1.1% | 1.00 |
| Vision loss (%) |  | 1.2% | 1.2% | 1.00 |
| Lifestyle |  |  |  |  |
| Physical Activity | Moderate | 10.91 (10.64) | 11.53 (11.17) | 0.14 |
|  | Vigorous | 3.06 (5.37) | 3.58 (6.20) | 0.01* |
| Smoker Status (% ever smoker) |  | 48.4% | 50.8% | 0.20 |
| Jenkins sleep problem score |  | 9.69 (4.47) | 9.73 (4.29) | 0.82 |
| Fruit and vegetable intake (%) | Seldom | 0.1% | 0.4% | <0.001*** |
|  | < 1 per month | 0.3% | 0.2% |  |
|  | 1-3 per month | 0.7% | 0.4% |  |
|  | 1-2 per week | 2.5% | 1.1% |  |
|  | 3-4 per week | 7.2% | 5.5% |  |
|  | 5-6 per week | 8.5% | 6.5% |  |
|  | 1 per day | 20.3% | 17.3% |  |
|  | 2-3 daily | 45.1% | 47.8% |  |
|  | 4+ daily | 15.3% | 21.0% |  |
| Alcoholic units per week |  | 9.04 (10.16) | 9.76 (11.02) | 0.06 |
| Social activity engagement | Social activities | 13.32 (3.23) | 13.94 (3.16) | <0.001*** |
|  | Individualistic activities | 18.56 (3.30) | 19.17 (3.19) | <0.001*** |
| *Psychosocial* |  |  |  |  |
| CES-D Score |  | 6.33 (6.87) | 5.71 (6.43) | <0.001*** |

*N.b.* CN Cognitively normal, SE standard error, M mean, \* $p < 0.05$ , \*\* $p < 0.01$ , \*\*\* $p <$ 0.001

### Binary logistic regression results

The adjusted binary logistic regression analysis is presented in Table 3. Examining the demographic factors in the model, we observed that each increase in education level was associated with a 23% increase in the odds of being a SuperAger (OR = 1.23, SE = 0.06, 95% CI = [1.11, 1.38], *p* < 0.001). In addition, increasing age group was associated with a 33% reduction in the odds of being a SuperAger (OR = 0.67, SE = 0.04, 95% CI = [0.61, 0.72], *p* < 0.001). Sex was also significantly associated with SuperAger status, with females having 30% higher odds of being a SuperAger (OR = 1.30, SE = 0.09, 95% CI = [1.10, 1.55], *p* = 0.003).

**Table 3.** Binary logistic regression results of different dementia risk factors on SuperAger status.

| Variable | OR | SE | 95% CI | $p$ value |
| --- | --- | --- | --- | --- |
| *Demographic* |  |  |  |  |
| **Education level<sup>^</sup>** | **1.23** | **0.06** | **[1.11, 1.38]** | **<0.001\*\*\*** |
| **Age group<sup>^</sup>** | **0.67** | **0.04** | **[0.61, 0.72]** | **<0.001\*\*\*** |
| Father's social class<sup>^</sup> | 1.00 | 0.04 | [0.93, 1.07] | 0.95 |
| **Sex** | **1.30** | **0.09** | **[1.10, 1.55]** | **0.003\*\*** |
| *Biomedical* |  |  |  |  |
| Total BMI | 0.99 | 0.01 | [0.97, 1.00] | 0.15 |
| Total medications | 1.01 | 0.02 | [0.98, 1.04] | 0.64 |
| **Hypertension** | **0.77** | **0.08** | **[0.66, 0.91]** | **0.002\*\*** |
| Diabetes | 0.91 | 0.15 | [0.69, 1.22] | 0.54 |
| *Lifestyle* |  |  |  |  |
| Moderate/vigorous exercise | 1.00 | 0.00 | [0.99, 1.01] | 0.86 |
| **Social activity engagement** | **1.03** | **0.01** | **[1.01, 1.06]** | **0.02\*** |
| Individualised activity engagement | 1.02 | 0.01 | [0.99, 1.05] | 0.12 |
| **Fruit and vegetable intake** | **1.08** | **0.03** | **[1.02, 1.15]** | **0.01\*** |
| *Psychosocial* |  |  |  |  |
| **CES-D score** | **0.99** | **0.01** | **[0.97, 1.00]** | **0.03\*** |

N.b. \* $p < 0.05$ , \*\* $p < 0.01$ , \*\*\* $p < 0.001$ , CI confidence interval, OR odd ratios, SE standard error, <sup>^</sup> treated as categorical then numeric for reporting

Examining dementia risk factors, several were associated with SuperAger status. Hypertension was associated with a 23% reduction in the odds of being a SuperAger (OR = 0.77, SE = 0.08, 95% CI = [0.66, 0.91], *p* = 0.002). Higher CES-D score was associated with reduced odds of being a SuperAger, with each one-point increase in CES-D score decreasing the odds of being a SuperAger by approximately 1% (OR = 0.99, SE = 0.01, 95% CI = [0.97, 1.00], *p* = 0.03). Higher engagement in social activities was associated with increased odds of being a SuperAger, with each unit increase in social activity engagement associated with a 3% increase in the odds of being a SuperAger (OR = 1.03, SE = 0.01, 95% CI = [1.01, 1.06], *p* = 0.02). Higher fruit and vegetable intake was associated with increased odds of being a SuperAger, with each unit increase on the fruit and vegetable scale associated with an 8% increase in the odds of being a SuperAger (OR = 1.08, SE = 0.03, 95% CI = [1.02, 1.15], *p* = 0.01). None of the other examined factors were observed to relate to SuperAger status (see Table 3).

## Discussion

In this large population study of British civil servants followed into older age we set out to investigate the relationship between established dementia risk factors and the odds of being a SuperAger. We observed higher education level was associated with an increase in the odds of being a SuperAger, while higher age group also associated was associated with reduced odds of being a SuperAger. Hypertension and higher depression score were associated with a reduction in the odds of being a SuperAger, while higher social activity engagement and higher fruit and vegetable intake were associated with an increase in the odds of being a SuperAger. Several risk factors previously observed to be associated with dementia were not significant in the present study, including BMI, physical activity, smoking, and alcohol consumption.

The finding that SuperAgers reported higher levels of social engagement aligns with literature on cognitive ageing and dementia. For instance, although not specifically examining SuperAgers, James and colleagues’ (2011) study of late-life social activity found that higher social activity scores were associated with reduced cognitive decline in older age (James et al., 2011). Sensitivity analyses further indicated that this association was not driven by individuals with low baseline cognition or mild cognitive impairment, suggesting that greater social engagement is linked to better cognitive ageing in their analysis. Moreover, in Saint Martin and colleagues’ (2017) study investigating the role of lifestyle factors in cognitive elites (CEs), they observed greater participation in social activities in CEs, with stability in cognitive ability. However, based on the findings of this study, we cannot be certain of the directionality of this relationship; that is, whether increasing engagement in social activities increases the likelihood of becoming a SuperAger, or whether SuperAgers show higher engagement in social activities. Moreover, it is important to acknowledge the effect size was modest, and as such, caution should be taken to not overinterpret the potential impact of social engagement.

The present studies reporting of an association between diet (fruit and vegetable intake) and SuperAger status is also broadly consistent with the limited literature examining diet in SuperAgers. Wagner and Grodstein (2022) similarly reported that female exceptional memory agers demonstrated greater adherence to the Mediterranean diet during both midlife and later life than cognitively normal controls, suggesting that healthier dietary patterns may contribute to exceptional cognitive ageing (Wagner & Grodstein, 2022). In contrast, the only other study to investigate diet in SuperAgers did not observe a significant association, although this may reflect its relatively small sample size (64 SuperAgers and 55 cognitively normal participants), limiting statistical power. Given the scarcity of SuperAger research, evidence from studies of cognitive ageing provides important context. Consistent with the present findings, reviews have concluded that greater fruit and vegetable intake is generally associated with better cognitive function and slower cognitive decline in older adults (Haskell-Ramsay & Docherty, 2023; Loef & Walach, 2012). Together, these findings suggest that the association observed in the present study extends existing evidence by demonstrating that greater fruit and vegetable intake is also associated with the preservation of exceptional memory performance in older age. Future studies should investigate whether this relationship is driven by specific dietary components, such as vegetable intake, or broader dietary patterns, including the Mediterranean or MIND diets.

The present study observed lower depression scores to be a significant predictor of SuperAger status. The association between depression, dementia risk and cognitive decline is well established (Fernández Fernández et al., 2024; Zheng et al., 2018), with these findings suggesting that psychological wellbeing may play an important role in maintaining cognitive function, rather than solely preventing cognitive decline. This is consistent with findings from several SuperAging studies (Doyle et al., 2024; Hermansen et al., 2024; Maccora et al., 2021; Powell et al., 2026). For instance, Doyle and colleagues (2024) identified lower depression symptoms as a feature of SuperAging in two Hispanic samples. While some SuperAging studies have not observed significant associations, these tended to include smaller sample sizes (Cook Maher et al., 2022; Dang et al., 2019), which may have limited statistical power. Taken together, these findings suggest that lower levels of depressive symptoms may represent a consistent characteristic of SuperAging, highlighting the importance of psychological wellbeing in exceptional cognitive ageing.

The observed association between hypertension and reduced odds of SuperAger status is consistent with a substantial body of research linking hypertension to increased dementia risk. Studies have shown that hypertension is associated with a higher risk of all-cause dementia (Livingston et al., 2024; Sharp et al., 2011), with evidence also indicating that greater variability in systolic blood pressure is associated with an increased risk of dementia (Mahinrad et al., 2023). These associations are thought to operate through several biological mechanisms, including cerebral small vessel disease, impaired cerebral perfusion, and disruption of the blood–brain barrier, which together contribute to neuronal damage and cognitive decline (Canavan & O’Donnell, 2022). In this context, SuperAgers may be less exposed to these vascular and neurodegenerative processes or may be more resilient to their effects, contributing to cognitive preservation. However, findings regarding hypertension and SuperAger status are inconsistent. While Garo-Pascual and colleagues (2023) and Powell and colleagues (2025) identified hypertension as a significant predictor of SuperAger status (Garo-Pascual et al., 2023; Powell et al., 2026), some studies failed to report a significant association (Calandri et al., 2020; Dang et al., 2019; Doyle et al., 2024). These discrepancies may reflect differences in sample characteristics, measurement of hypertension, or levels of treatment and control across cohorts, highlighting the need for further research to clarify this relationship.

Overall, the present study observed several modest but promising results on the relationship between late-life risk factors and SuperAger status. These modest effect sizes however while indicating important distinctions between SuperAgers and controls, suggest that these factors are unlikely to be the primary determinants of exceptional cognitive ageing. This raises the question of what mechanisms underlie SuperAging. One possibility is that the influence of lifestyle on cognition operates during earlier, sensitive periods of life, underscoring the importance of a life-course perspective.

Consistent with this, Gow and colleagues (2017), using the Lothian Birth Cohort 1921, found that childhood ability and engagement in activities of a social or intellectual nature during mid-life were associated with higher cognitive ability, while later-life activities were rather associated with level of decline in cognitive abilities (Gow et al., 2017). Similar patterns were observed in Richards and colleagues (2003) study on older-middle aged adults using the National Survey for Health and Development (NSHD) 1946 birth cohort (Richards et al., 2003). Future research should, therefore, adopt a longitudinal, life-course approach to better understand how and whether life course factors contribute to exceptional cognitive ageing.

While a life-course perspective may explain the limited impact of late-life risk factors, an alternative explanation for the limited impact of these factors, is that SuperAger status may be strongly influenced by non-modifiable factors, such as genetics and early-life cognitive ability. Supporting this, studies demonstrate remarkable stability in cognitive ability across the lifespan (Gow et al., 2011; Nyberg & Pudas, 2019). For example, the Lothian Birth Cohorts reported high stability coefficients for cognitive performance from age 11 to age 70 (r = 0.67) and from age 11 to age 87 (r = 0.51) (Gow et al., 2011; Nyberg & Pudas, 2019). Similarly, findings from the Swedish Betula cohort indicate strong stability in general cognitive ability from early adulthood (age 18) into later life (age 50–65) (Nyberg & Pudas, 2019; Rönnlund et al., 2015).

While the present studies strengths include the large sample size and comparison of several risk factors with SuperAger status, several limitations are important to acknowledge. Firstly, the Whitehall II cohort, consisting predominantly of white-collar workers in stable employment (two-thirds men), lacks representativeness of the general population, thus, the generalisability of our findings requires investigation in more diverse cohorts. Second, our SuperAgers were relatively young (mostly under 80) compared to those typically studied in the SuperAger literature (often over 80). Finally, the cross-sectional nature of our analysis precludes directional knowledge and causal inference.

## Conclusion

Overall, this study found that several late-life risk factors, including depressive symptoms, absence of hypertension, higher fruit and vegetable intake and higher social engagement, were associated with SuperAger status. However, the modest effect sizes observed suggest that late-life risk factors alone are unlikely to fully explain SuperAging. Future research would therefore benefit from examining risk factors longitudinally across the lifespan, as well as the potential role of genetic and other non-modifiable factors, to better understand the mechanisms underlying SuperAging.

## Author contributions

Conceptualization: Philippa Watson; Data analysis: Philippa Watson; Writing - original draft preparation: Philippa Watson; Writing - review and editing: Sarah Bauermeister, Omid Ebrahimi, John Gallacher; Supervision: Sarah Bauermeister, Omid Ebrahimi

## Data Availability

All data is freely available via application through Dementias Platform UK.

## References

Ainsworth, B. E., Haskell, W. L., Whitt, M. C., Irwin, M. L., Swartz, A. M., Strath, S. J., O Brien, W. L., Bassett, D. R., Schmitz, K. H., & Emplaincourt, P. O. (2000). Compendium of physical activities: an update of activity codes and MET intensities. Medicine and science in sports and exercise, 32(9; SUPP/1), S498-S504.

Anstey, K., & Christensen, H. (2000). Education, activity, health, blood pressure and apolipoprotein E as predictors of cognitive change in old age: a review. Gerontology, 46(3), 163–177. 10.1159/000022153

Baumgart, M., Snyder, H. M., Carrillo, M. C., Fazio, S., Kim, H., & Johns, H. (2015). Summary of the evidence on modifiable risk factors for cognitive decline and dementia: a population-based perspective. Alzheimer’s & Dementia, 11(6), 718–726.

Borkowski, J. G., Benton, A. L., & Spreen, O. (1967). Word fluency and brain damage. Neuropsychologia, 5(2), 135–140.

Burke, S. N., Mormino, E. C., Rogalski, E. J., Kawas, C. H., Willis, R. J., & Park, D. C. (2019). What are the later life contributions to reserve, resilience, and compensation? Neurobiology of aging, 83, 140–144.

Calandri, I. L., Crivelli, L., Martin, M. E., Egido, N., Guimet, N. M., & Allegri, R. F. (2020). Environmental factors between normal and superagers in an Argentine cohort. Dementia & Neuropsychologia, 14, 345–349.

Canavan, M., & O’Donnell, M. J. (2022). Hypertension and cognitive impairment: a review of mechanisms and key concepts. Frontiers in Neurology, 13, 821135.

Cook Maher, A., Makowski-Woidan, B., Kuang, A., Zhang, H., Weintraub, S., Mesulam, M. M., & Rogalski, E. (2022). Neuropsychological Profiles of Older Adults with Superior versus Average Episodic Memory: The Northwestern “SuperAger” Cohort. J Int Neuropsychol Soc, 28(6), 563–573. 10.1017/S1355617721000837

Dang, C., Harrington, K. D., Lim, Y. Y., Ames, D., Hassenstab, J., Laws, S. M., Yassi, N., Hickey, M., Rainey-Smith, S. R., Robertson, J., Rowe, C. C., Sohrabi, H. R., Salvado, O., Weinborn, M., Villemagne, V. L., Masters, C. L., Maruff, P., & Group, A. R. (2019). Superior Memory Reduces 8-year Risk of Mild Cognitive Impairment and Dementia But Not Amyloid beta-Associated Cognitive Decline in Older Adults. Arch Clin Neuropsychol, 34(5), 585–598. 10.1093/arclin/acy078

de Godoy, L. L., Alves, C., Saavedra, J. S. M., Studart-Neto, A., Nitrini, R., da Costa Leite, C., & Bisdas, S. (2021). Understanding brain resilience in superagers: a systematic review. Neuroradiology, 63(5), 663–683. 10.1007/s00234-020-02562-1

Dekhtyar, M., Papp, K. V., Buckley, R., Jacobs, H. I. L., Schultz, A. P., Johnson, K. A., Sperling, R. A., & Rentz, D. M. (2017). Neuroimaging markers associated with maintenance of optimal memory performance in late-life. Neuropsychologia, 100, 164–170. 10.1016/j.neuropsychologia.2017.04.037

Dintica, C. S., & Yaffe, K. (2022). Epidemiology and risk factors for dementia. Psychiatric Clinics, 45(4), 677–689.

Doyle, C., Andel, R., Saenz, J., & Crowe, M. (2024). Correlates of SuperAging in Two Population-Based Samples of Hispanic Older Adults. J Gerontol B Psychol Sci Soc Sci, 79(6). 10.1093/geronb/gbae058

Fernández Fernández, R., Martín, J. I., & Antón, M. A. M. (2024). Depression as a risk factor for dementia: a meta-analysis. The Journal of Neuropsychiatry and Clinical Neurosciences, 36(2), 101–109.

Fotenos, A. F., Snyder, A. Z., Girton, L. E., Morris, J. C., & Buckner, R. L. (2005). Normative estimates of cross-sectional and longitudinal brain volume decline in aging and AD. Neurology, 64(6), 1032–1039. 10.1212/01.WNL.0000154530.72969.11

Garo-Pascual, M., Gaser, C., Zhang, L., Tohka, J., Medina, M., & Strange, B. A. (2023). Brain structure and phenotypic profile of superagers compared with age-matched older adults: a longitudinal analysis from the Vallecas Project. Lancet Healthy Longev, 4(8), e374–e385. 10.1016/S2666-7568(23)00079-X

Gefen, T., Peterson, M., Papastefan, S. T., Martersteck, A., Whitney, K., Rademaker, A., Bigio, E. H., Weintraub, S., Rogalski, E., Mesulam, M. M., & Geula, C. (2015). Morphometric and histologic substrates of cingulate integrity in elders with exceptional memory capacity. J Neurosci, 35(4), 1781–1791. 10.1523/JNEUROSCI.2998-14.2015

Gow, A. J., Johnson, W., Pattie, A., Brett, C. E., Roberts, B., Starr, J. M., & Deary, I. J. (2011). Stability and change in intelligence from age 11 to ages 70, 79, and 87: the Lothian Birth Cohorts of 1921 and 1936. Psychology and aging, 26(1), 232.

Gow, A. J., Pattie, A., & Deary, I. J. (2017). Lifecourse Activity Participation From Early, Mid, and Later Adulthood as Determinants of Cognitive Aging: The Lothian Birth Cohort 1921. J Gerontol B Psychol Sci Soc Sci, 72(1), 25–37. 10.1093/geronb/gbw124

Harrison, T. M., Maass, A., Baker, S. L., & Jagust, W. J. (2018). Brain morphology, cognition, and beta-amyloid in older adults with superior memory performance. Neurobiol Aging, 67, 162–170. 10.1016/j.neurobiolaging.2018.03.024

Harrison, T. M., Weintraub, S., Mesulam, M. M., & Rogalski, E. (2012). Superior memory and higher cortical volumes in unusually successful cognitive aging. J Int Neuropsychol Soc, 18(6), 1081–1085. 10.1017/S1355617712000847

Haskell-Ramsay, C. F., & Docherty, S. (2023). Role of fruit and vegetables in sustaining healthy cognitive function: evidence and issues. Proceedings of the Nutrition Society, 82(3), 305–314.

Hermansen, M., Nygaard, M., Tan, Q., Jeune, B., Semkovska, M., Christensen, K., Thinggaard, M., & Mengel-From, J. (2024). Cognitively high-performing oldest old individuals are physically active and have strong motor skills-A study of the Danish 1905 and 1915 birth cohorts. Arch Gerontol Geriatr, 122, 105398. 10.1016/j.archger.2024.105398

James, B. D., Wilson, R. S., Barnes, L. L., & Bennett, D. A. (2011). Late-life social activity and cognitive decline in old age. J Int Neuropsychol Soc, 17(6), 998–1005. 10.1017/S1355617711000531

Jenkins, C. D., Stanton, B. A., Niemcryk, S. J., & Rose, R. M. (1988). A scale for the estimation of sleep problems in clinical research. J Clin Epidemiol, 41(4), 313–321. 10.1016/0895-4356(88)90138-2

Livingston, G., Huntley, J., Liu, K. Y., Costafreda, S. G., Selbaek, G., Alladi, S., Ames, D., Banerjee, S., Burns, A., Brayne, C., Fox, N. C., Ferri, C. P., Gitlin, L. N., Howard, R., Kales, H. C., Kivimaki, M., Larson, E. B., Nakasujja, N., Rockwood, K.,…Mukadam, N. (2024). Dementia prevention, intervention, and care: 2024 report of the Lancet standing Commission. Lancet, 404(10452), 572–628. 10.1016/S0140-6736(24)01296-0

Loef, M., & Walach, H. (2012). Fruit, vegetables and prevention of cognitive decline or dementia: a systematic review of cohort studies. The Journal of nutrition, health and aging, 16(7), 626–630.

Lourida, I., Hannon, E., Littlejohns, T. J., Langa, K. M., Hyppönen, E., Kuźma, E., & Llewellyn, D. J. (2019). Association of lifestyle and genetic risk with incidence of dementia. JAMA, 322(5), 430–437.

Maccora, J., Peters, R., & Anstey, K. J. (2021). Gender Differences in Superior-memory SuperAgers and Associated Factors in an Australian Cohort. J Appl Gerontol, 40(4), 433–442. 10.1177/0733464820902943

Mahinrad, S., Bennett, D. A., Sorond, F. A., & Gorelick, P. B. (2023). Blood pressure variability, dementia, and role of antihypertensive medications in older adults. Alzheimer’s & Dementia, 19(7), 2966–2974.

Marmot, M., & Brunner, E. (2005). Cohort Profile: the Whitehall II study. Int J Epidemiol, 34(2), 251–256. 10.1093/ije/dyh372

Martin, P., Kelly, N., Kahana, B., Kahana, E., Willcox, B. J., Willcox, D. C., & Poon, L. W. (2015). Defining Successful Aging: A Tangible or Elusive Concept? The Gerontologist, 55(1), 14–25. 10.1093/geront/gnu044

Mukadam, N., Anderson, R., Knapp, M., Wittenberg, R., Karagiannidou, M., Costafreda, S. G., Tutton, M., Alessi, C., & Livingston, G. (2020). Effective interventions for potentially modifiable risk factors for late-onset dementia: a costs and cost-effectiveness modelling study. Lancet Healthy Longev, 1(1), e13–e20. 10.1016/S2666-7568(20)30004-0

Mukadam, N., Wolters, F. J., Walsh, S., Wallace, L., Brayne, C., Matthews, F. E., Sacuiu, S., Skoog, I., Seshadri, S., & Beiser, A. (2024). Changes in prevalence and incidence of dementia and risk factors for dementia: an analysis from cohort studies. The Lancet Public Health, 9(7), e443–e460.

Nyberg, L., & Pudas, S. (2019). Successful memory aging. Annual review of psychology, 70(1), 219–243.

Peters, R., Booth, A., Rockwood, K., Peters, J., D’Este, C., & Anstey, K. J. (2019). Combining modifiable risk factors and risk of dementia: a systematic review and meta-analysis. BMJ Open, 9(1), e022846. 10.1136/bmjopen-2018-022846

Petersen, R. C., Caracciolo, B., Brayne, C., Gauthier, S., Jelic, V., & Fratiglioni, L. (2014). Mild cognitive impairment: a concept in evolution. J Intern Med, 275(3), 214–228. 10.1111/joim.12190

Powell, A., Humburg, P., Kochan, N. A., Crawford, J. D., Wen, W., Close, J. C., Sachdev, P. S., & Brodaty, H. (2026). Characteristics of Australian cognitive super-agers using different measurement approaches. Aging & Mental Health, 1–13.

Radloff, L. S. (1977). The CES-D scale: A self-report depression scale for research in the general population. Applied psychological measurement, 1(3), 385–401.

Resnick, S. M., Pham, D. L., Kraut, M. A., Zonderman, A. B., & Davatzikos, C. (2003). Longitudinal magnetic resonance imaging studies of older adults: a shrinking brain. J Neurosci, 23(8), 3295–3301. 10.1523/JNEUROSCI.23-08-03295.2003

Richards, M., Hardy, R., & Wadsworth, M. E. (2003). Does active leisure protect cognition? Evidence from a national birth cohort. Social science & medicine, 56(4), 785–792.

Richardson, M. T., Leon, A. S., Jacobs Jr, D. R., Ainsworth, B. E., & Serfass, R. (1994). Comprehensive evaluation of the Minnesota leisure time physical activity questionnaire. Journal of clinical epidemiology, 47(3), 271–281.

Rogalski, E. J., Gefen, T., Shi, J., Samimi, M., Bigio, E., Weintraub, S., Geula, C., & Mesulam, M.-M. (2013). Youthful memory capacity in old brains: anatomic and genetic clues from the Northwestern SuperAging Project. Journal of cognitive neuroscience, 25(1), 29–36.

Rönnlund, M., Sundström, A., & Nilsson, L.-G. (2015). Interindividual differences in general cognitive ability from age 18 to age 65 years are extremely stable and strongly associated with working memory capacity. Intelligence, 53, 59–64.

Rowe, J. W., & Kahn, R. L. (2015). Successful aging 2.0: Conceptual expansions for the 21st century. In (Vol. 70, pp. 593–596): Oxford University Press US.

Rubin, D. B. (2018). Multiple imputation. In Flexible imputation of missing data, second edition (pp. 29-62). Chapman and Hall/CRC.

Sabia, S., Dugravot, A., Dartigues, J. F., Abell, J., Elbaz, A., Kivimaki, M., & Singh-Manoux, A. (2017). Physical activity, cognitive decline, and risk of dementia: 28 year follow-up of Whitehall II cohort study. BMJ, 357, j2709. 10.1136/bmj.j2709

Sabia, S., Dugravot, A., Kivimaki, M., Brunner, E., Shipley, M. J., & Singh-Manoux, A. (2012). Effect of intensity and type of physical activity on mortality: results from the Whitehall II cohort study. Am J Public Health, 102(4), 698–704. 10.2105/AJPH.2011.300257

Sharp, S. I., Aarsland, D., Day, S., Sønnesyn, H., Group, A. s. S. V. D. S. R., & Ballard, C. (2011). Hypertension is a potential risk factor for vascular dementia: systematic review. International Journal of Geriatric Psychiatry, 26(7), 661–669.

Singh-Manoux, A., Richards, M., & Marmot, M. (2003). Leisure activities and cognitive function in middle age: evidence from the Whitehall II study. J Epidemiol Community Health, 57(11), 907–913. 10.1136/jech.57.11.907

Statistics, O. f. N. (2024). Population estimates for the UK, England, Wales, Scotland, and Northern Ireland: mid-2022. Retrieved Aug. 12 from

Sun, F. W., Stepanovic, M. R., Andreano, J., Barrett, L. F., Touroutoglou, A., & Dickerson, B. C. (2016). Youthful Brains in Older Adults: Preserved Neuroanatomy in the Default Mode and Salience Networks Contributes to Youthful Memory in Superaging. J Neurosci, 36(37), 9659–9668. 10.1523/JNEUROSCI.1492-16.2016

Wagner, M., & Grodstein, F. (2022). Patterns of lifestyle behaviours from mid-through later-life in relation to exceptional episodic memory performance in older women: the Nurses’ Health Study. Age Ageing, 51(5). 10.1093/ageing/afac102

White, I. R., Royston, P., & Wood, A. M. (2011). Multiple imputation using chained equations: issues and guidance for practice. Statistics in medicine, 30(4), 377–399.

Yang, Z., Wen, W., Jiang, J., Crawford, J. D., Reppermund, S., Levitan, C., Slavin, M. J., Kochan, N. A., Richmond, R. L., Brodaty, H., Trollor, J. N., & Sachdev, P. S. (2016). Age-associated differences on structural brain MRI in nondemented individuals from 71 to 103 years. Neurobiol Aging, 40, 86–97. 10.1016/j.neurobiolaging.2016.01.006

Ye, K. X., Sun, L., Wang, L., Khoo, A. L. Y., Lim, K. X., Lu, G., Yu, L., Li, C., Maier, A. B., & Feng, L. (2023). The role of lifestyle factors in cognitive health and dementia in oldest-old: A systematic review. Neurosci Biobehav Rev, 152, 105286. 10.1016/j.neubiorev.2023.105286

Yu, J., Collinson, S. L., Liew, T. M., Ng, T. P., Mahendran, R., Kua, E. H., & Feng, L. (2020). Super-cognition in aging: Cognitive profiles and associated lifestyle factors. Appl Neuropsychol Adult, 27(6), 497–503. 10.1080/23279095.2019.1570928

Zheng, F., Zhong, B., Song, X., & Xie, W. (2018). Persistent depressive symptoms and cognitive decline in older adults. The British Journal of Psychiatry, 213(5), 638–644.

